# AI Identification of Transdiagnostic Factors in Adolescent Mental Health

**DOI:** 10.64898/2026.11.20.26368571

**Authors:** Lei Luo, Nicholas Cummins, Giorgia Michelini, Toby Wise, Raquel Iniesta

## Abstract

**Background:** Adolescent psychopathology is commonly assessed using high-dimensional questionnaire data. These data often consist of ordinal, sparse, and overlapping symptom items, creating challenges for deriving latent phenotypes that are both interpretable and generalisable. This study addressed the need to derive low- dimensional transdiagnostic phenotypes from adolescent mental health data by evaluating denoising autoencoders as a flexible non-linear representation-learning approach.

**Methods:** Using baseline data from the Adolescent Brain Cognitive Development Study, we analysed mental health questionnaire data from a large community sample of adolescents (N = 9,799). A denoising autoencoder was trained to learn low- dimensional latent representations and was benchmarked against principal component analysis, exploratory factor analysis, and radial basis function kernel principal component analysis using held-out reconstruction explained variance. Interpretability was assessed using item-level canonical correlation analyses and alignment with a framework of well-established psychopathology dimensions. External validity was examined using gradient-boosted tree models predicting clinical and functional validators.

**Results:** Across three to five factors solutions, the autoencoder consistently achieved the highest reconstruction explained variance compared with all benchmark methods. The five-factor autoencoder solution showed interpretable correspondence with established dimensional structures of psychopathology, rather than mapping onto a single general factor. Autoencoder-derived factors showed equal or modestly superior predictive validity compared with EFA across most validator domains.

**Conclusions:** These findings suggest that denoising autoencoders may offer a useful complementary approach for deriving low-dimensional, interpretable, and externally valid transdiagnostic phenotypes from adolescent symptom data, with potential utility for summarising complex symptom profiles and supporting dimensional assessment in clinical and research settings.

## Introduction

Most mental disorders begin before adulthood, making preadolescence and early adolescence a significant period for identifying at-risk phenotypes (Solmi et al., 2022). At the same time, youth psychopathological disorders are usually comorbid, thus any attempts to link one diagnostic label to aetiology or neurobiology have been challenging (Caspi et al., 2014; Kieling et al., 2011; Lahey et al., 2021). A central challenge is therefore how to characterise emotional and behavioural problems, which are typically measured using many overlapping symptoms rather than a single summary score, in a lower-dimensional and analytically useful form.

Current frameworks increasingly emphasise dimensional and transdiagnostic descriptions of mental health disorders. These frameworks integrate symptoms into broader domains to better match the structure observed in data and account for comorbidity and overlap among symptoms. In doing so, these approaches aim to facilitate research by avoiding a focus on diagnostic categories which are likely not underpinned by clearly distinct pathology (Insel et al., 2010; Kotov et al., 2017). At a methodological level, dimensionality reduction methods such as exploratory factor analysis (EFA) have been used to find transdiagnostic factors (Michelini et al., 2019). EFA can transform high dimensional symptoms into lower dimensional latent phenotypes that allows to capture shared features, reduce redundancy, and support downstream mechanistic interpretability (Conway et al., 2019). Prior work has shown that childhood psychopathology can be organised into higher-order dimensions with meaningful external correlates using linear methods (Holmes et al., 2021; Michelini et al., 2019; Modabbernia et al., 2022). For example, Michelini et al. (2019) used hierarchical EFA to identify a hierarchy of dimensions in the ABCD study sample, showing meaningful associations with clinically relevant validators at different levels of this hierarchy.

Despite the value of dimensionality reduction methods, measures of youth psychopathology based on questionnaires pose several challenges for dimensionality reduction. First, item responses are often sparse, and skewed in many samples, which can mislead downstream factor models (Rhemtulla et al., 2012). Second, item sets often contain substantial redundancy, including highly similar or tightly coupled content that can complicate the recovery of a stable latent structure (Fried, 2017). Although redundancy is commonly addressed by dropping or collapsing items, such action can make latent solutions sensitive to preprocessing decisions. Third, dimensionality reduction also raises a core evaluation problem: the reduced dimension space should be stable, reproducible, or useful outside the sample from which they were derived. Nested cross-validation has long been recommended, because it separates hyperparameter search from held-out evaluation and reduces optimism in error estimates (Varma & Simon, 2006). This highlights the need to assess robustness across repeated runs, rather than relying on a single train-test split or a single cross- validation run (Bouthillier et al., 2021; D’Amour et al., 2022).

Linear approaches have the limitations that they impose structural assumptions that may be restrictive when symptom relationships are shaped by thresholds, interactions, or other non-linear dependencies (Hinton & Salakhutdinov, 2006). Non-linear approaches therefore warrant explicit evaluation in youth mental health data, where questionnaire responses are often sparse, ordinal, and potentially heterogeneous in structure. Non-linear approaches such as kernel principal component analysis (kernel PCA) aim to capture curved or manifold structure by implicitly mapping inputs into a high dimensional feature space before performing PCA. It offers a flexible alternative when relationships are not well approximated by linear subspaces (Schölkopf et al., 1998). These approaches have different inductive biases, and in practice they may trade interpretability, robustness, and sensitivity to preprocessing decisions in different ways, providing essential benchmarking for mental health data dimension reduction.

Autoencoders provide a flexible representation learning approach to derive low- dimensional latent phenotypes from high-dimensional symptom data, with the aim of capturing potentially non-linear symptom relationships while reducing redundancy and preserving clinically meaningful variation (Hinton & Salakhutdinov, 2006; Milano et al., 2024; Ostojic et al., 2024). They consist of two parts: an encoder, which compresses the original input into a lower-dimensional latent space, and a decoder, which reconstructs the input from this compressed representation. Because these models are trained to preserve the most informative patterns needed for reconstruction, the learned latent representation can be used as a data-driven phenotype embedding (Hinton & Salakhutdinov, 2006). As a class of neural networks, autoencoders can represent nonlinear functions and are not restricted to orthogonal components or linear factor structure, potentially capturing patterns in symptom profiles that arise from interactions, thresholds, or other nonlinearities (Bengio et al., 2013).

In this study, we evaluate a novel, non-linear dimensionality-reduction pipeline for youth mental health phenotyping from survey items. Building on the dimensionality reduction and evaluation framework established by Michelini et al. (2019), we examine whether non-linear representation learning can derive low-dimensional phenotypes that are reconstructive and interpretable and benchmark this approach against PCA, EFA, and kernel PCA. Finally, we examine interpretability using canonical-correlation analysis (CCA)-based analyses to relate the learned phenotypes to the original survey items. Our results indicate that nonlinear representation learning can support the construction of reproducible low-dimensional phenotypes for developmental mental health research.

## Methods

### Participants

This study used data from the Adolescent Brain Cognitive Development (ABCD) Study, an ongoing multisite longitudinal cohort in the United States that recruited approximately 11,500 children through school systems at 21 research sites (Garavan et al., 2018). Participants were aged 9–10 years at the baseline assessment from 2016 to 2018 and are planned to be followed for approximately 10 years into early adulthood. The ABCD protocol includes harmonized neuroimaging acquisition across the 21 sites (Casey et al., 2018). Data were accessed via the NIMH Data Archive (NDA) using the ABCD Curated Annual Release 5.1 (DOI: 10.15154/z563-zd24).

ABCD data collection procedures were reviewed and approved by a central institutional review board and, when applicable, local review boards, with written informed consent from caregivers and assent from children. Secondary use of the ABCD data complied with NDA data use requirements, and the present analyses received approval from an NIH Data Access Committee.

### Measures

Psychopathology symptoms were assessed using the Child Behaviour Checklist (CBCL), a widely used dimensional measure of emotional and behavioural problems in children and adolescents (Achenbach & Ruffle, 2000). Parents rated symptom items at baseline using the standard three-point CBCL response format, with higher scores indicating greater symptom endorsement. For the present analyses, we extracted CBCL data and validator data collected at baseline. Item-level CBCL data were used as the input symptom features for dimensionality reduction analyses.

Baseline validator data were extracted to assess the external validity of the derived latent phenotypes across clinical, cognitive, family, school, developmental, and peer-related domains. To reduce familial aggregation and the non-independence arising from shared genetic and environmental factors in households with multiple enrolled children, we retained only one randomly selected child per family, after dropping missing data, yielding a final analytic sample of 9799 out of 11862.

### Preprocessing

The measurement properties of questionnaire data require additional preprocessing of input data. Because questionnaire items are ordinal, association measures that explicitly model ordinality can be preferable to simple Pearson correlations. In practice, ordinal estimation strategies and the use of polychoric correlations have been shown to affect recovered dimensional structure, particularly when items have few categories or when distributions are skewed (Holgado–Tello et al., 2010; Rhemtulla et al., 2012). At the same time, high dimensional symptoms data often contain redundant items such as near duplicates or tightly coupled content clusters, which can complicate factor models.

Preprocessing was performed prior to model fitting. To reduce sparsity and to support ordinal association modelling, each CBCL item was screened for extremely low response frequency. Items were excluded if more than 99.5% of observed non-missing responses were “No”. All remaining items were treated as ordered categorical variables. A single polychoric correlation matrix was then estimated using pairwise handling of missing data, such that each correlation was computed from participants with non-missing responses on the corresponding item pair.

To reduce redundancy among retained items, a graph clustering approach was applied. A thresholded polychoric correlation matrix was constructed, in which items were connected when their pairwise association exceeded r > 0.75. Connected components were identified and each component was aggregated into a composite variable. For each participant, the composite value was computed as the mean of the component’s item responses and was then rounded to the nearest integer. The original items contributing to each composite were removed, and the derived composites were appended to the dataset.

### Benchmarking pipeline

A benchmarking pipeline was implemented to learn low dimensional latent scores from the CBCL feature matrix. We compared autoencoder with PCA, EFA, and kernel PCA using reconstruction performance evaluated on held out data. Analysis was done using Python v3.11.6, NumPy v1.23.5, and PyTorch v2.2.1. To promote reproducibility, we fixed random seeds across all packages, including CUDA where available, constrained thread usage, and enabled deterministic computation settings for the autoencoder where supported.

For autoencoder training and evaluation, the dataset was split into train, validation, and test partitions (80%/10%/10%). Within the training set, a nested cross-validation framework was employed for hyperparameter tuning and model selection. Specifically, the training data were further divided into outer and inner folds. The inner loop was used to optimise hyperparameters, while the outer loop provided an unbiased estimate of model performance during model development. The final model, with optimised hyperparameters, was retrained on the full training data and evaluated on the independent test set.

To minimise data leakage, feature standardisation was always estimated using training data only and then applied unchanged to validation and test data. Features were z-scored based on the distribution of the training partition. Within cross-validation, this procedure was implemented separately in each fold using scikit-learn pipelines, ensuring that fold-specific training statistics were not informed by held-out data.

Parallel analyses were used to guide the number of factors, and all models were evaluated at target latent dimensionalities from 1 to 10, chosen to reflect a parsimonious range consistent with prior dimensional work in child psychopathology and to enable fair comparison across methods. For each target latent dimensionality *d*, EFA, PCA and kernel PCA were fit on the standardised training data. Reconstructions were obtained via the corresponding inverse mappings for PCA and kernel PCA, whereas EFA reconstructions were computed in the standardised space by back-projecting factor scores through the rotated loading matrix. The reconstruction explained variance ratio (EVR) was computed as:

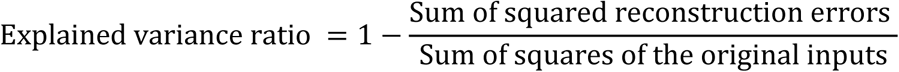

When necessary, hyperparameters were tuned with a randomised search strategy with cross-validation. Generalisation was assessed using a nested procedure with an outer 5-fold KFold, which separates model selection from performance estimation and reduces selection bias.

### Autoencoder Model

An autoencoder was implemented in PyTorch to learn a low dimensional latent representation from the input feature matrix X (tabular questionnaire or phenotype features) by minimizing reconstruction error using an encoder–decoder architecture (Hinton & Salakhutdinov, 2006). The autoencoder comprised a multilayer perceptron encoder with three fully connected (linear) layers that mapped the input to two hidden layers and then to a *d*-dimensional latent code. Gaussian error linear unit (GELU) nonlinearities were applied after the first two encoder layers. The decoder mirrored the encoder and used GELU nonlinearities after the first two decoder layers. The training objective was the MSE between the reconstructed output and the original uncorrupted inputs.

During training, denoising was induced via batch-swap noise, implemented by a feature-wise swapping of values across samples within each mini-batch with probability 0.1, while the reconstruction target remained the uncorrupted batch (Vincent et al., 2010). An optional decorrelation penalty was also tested. Within each mini-batch, the latent variables were mean-centred by subtracting the mean, and a covariance matrix was then computed from the centred latent representations to quantify dependence among latent dimensions, and the squared Frobenius norm of the off-diagonal elements was added to the loss with weight λ = 0.005 tuned over an empirically selected range from 0.001 to 0.01. The Adam optimizer was used with a learning rate of 0.003 and a L2 weight decay. Models were trained with a batch size of 32 for up to 1,000 epochs using a fixed training and validation split, with early stopping (patience = 10) based on validation. The learning rate was reduced by a factor of 10 when validation loss plateaued for 5 consecutive epochs. After training, latent factors, reconstructed inputs, reconstruction MSE per sample and EVR per factor were computed on held-out data.

#### Hyperparameters search strategy

Hidden-layer widths were searched over a discrete candidate set of 64 to 256 in increments of 32. We treated this architectural degree of freedom as a coarse design choice rather than an integer variable that required single-unit resolution. Restricting widths to meaningful discrete values reduced the effective dimensionality of the search space and improved the practicality of derivative-free optimization.

To identify a robust hyperparameter configuration for the final autoencoder model, we used 5-fold nested cross-validation. In each outer fold, hyperparameter optimization was performed using the corresponding training set and an inner validation procedure. Candidate configurations were proposed using a combination of the tree-structured Parzen estimator (TPE) sampler and covariance matrix adaptation evolution strategy (CMA-ES). The search procedure consisted of an initial broad search using TPE for 50 trials, followed by a more fine-grained optimisation using CMA-ES. Each candidate was evaluated using the prespecified reconstruction objective (EVR) computed on the inner validation data. This yielded one selected configuration per outer fold, together with its performance estimate on the held-out outer test partition.

Rather than selecting hyperparameters from the single outer fold with the highest outer test performance, which can introduce optimistic bias when test partitions influence model selection, we aggregated fold-wise selected hyperparameters across the five outer folds (Cawley & Talbot, 2010; Varma & Simon, 2006). For discrete architectural parameters, we selected the modal value across folds. For continuous hyperparameters, we summarized across folds using the median. This aggregation produced a final configuration intended to reflect stability across resamples rather than idiosyncrasies of a single split.

#### External Validation

To assess the external predictive validity of the latent representations, we replicated the analysis approach used by Michelini et al. (2019). Gradient boosting regression trees (GBRT) were used to model associations between latent factor scores and a set of external validators spanning clinical service use, medical history, medication use, family functioning, school-related measures, cognitive ability, developmental delays, and peer relationships. Clinical and health-related validators included parent-reported mental health service use, medical history, and medication use. Family and school validators included parent- and youth-reported measures of family functioning, school performance, and school-related difficulties. Cognitive validators were derived from baseline cognitive assessments, and developmental validators included parent-reported developmental delay indicators. Peer-related validators captured social functioning and peer relationship difficulties. These validators were not used during dimensionality reduction and were included only in downstream external validity analyses.

GBRT were trained to predict each validator from the latent factor scores derived using the autoencoder and EFA, with model performance quantified using total R^2^ (Nagelkerke R^2^ for binary outcomes). GBRT was chosen for its ability to capture non-linear relationships between predictor and target variables; given that the autoencoder method has the capacity to identify non-linear solutions, we may expect associations with external validators to also be non-linear. For each target dimensionality from 1 to 5, separate GBRT models were trained using latent factors derived from the autoencoder and EFA as predictors. For the GBRT models, we used the histogram-based gradient boosting implementation in scikit-learn. Specifically, the maximum number of boosting iterations was set to 100, corresponding to up to 100 trees; the learning rate was 0.10; the maximum number of leaf nodes per tree was 31; no explicit maximum tree depth was imposed; and L2 regularisation was set to 0. The minimum number of samples per leaf was 20. Hyperparameters were tuned using 5-fold cross-validation within the training data, and the optimised model was then evaluated on held-out data. Prediction performance was evaluated out of sample using the total coefficient of determination R^2^, with Nagelkerke R^2^ reported for binary validators. The same modelling framework and evaluation procedure were applied across all validators and representation types to ensure a fair comparison.

#### Interpretability

This study applied a multivariate association method, canonical correlation analysis (CCA), to seek linear combinations of latent space and original items that are maximally correlated, offering a natural framework for relating learned symptom phenotypes.

After hyperparameter selection, the final hyperparameter configuration was fixed and used to train the model repeatedly under multiple random seeds. For each run, we set the random seed across all packages and configured deterministic execution settings. Within each seed, model fitting was performed using 5-fold cross-validation. To obtain a single model instance for interpretability, we selected the fold whose mean reconstruction error was closest to the median across folds to avoid selecting an unusually favourable split, and we extracted latent factors for the CCA dataset using the trained encoder.

CCA was implemented between original items and latent factors. Before fitting CCA, both observed features and latent factors were standardized within the CCA routine using sample standard deviations. The number of canonical components was set to the same as latent factors. Canonical correlations were computed as the Pearson correlation between paired canonical variates. Permutation testing was performed with 1000 permutations during the process. Variable loadings were defined as correlations between standardized variables and canonical variates, and the top observed items per canonical component were summarized by absolute loading magnitude.

To examine the clinical interpretability of the autoencoder-derived latent factors, we evaluated their alignment with established Hierarchical Taxonomy of Psychopathology (HiTOP) spectra (Conway et al., 2019; Kotov et al., 2017). Each latent factor was represented by its item-level loading or contribution profile. To provide an indication of how these dimensions map onto existing taxonomies, we calculated an approximate mapping from our dimensions onto HiTOP symptom spectra. Alignment was quantified by mapping each item in the CBCL to the closest HiTOP dimension based on its content. For each autoencoder-derived latent factor, we then summed the weights for items within each HiTOP dimension. This analysis was used descriptively and approximately to assess whether the learned latent space captured differentiated and clinically interpretable symptom configurations.

To assess the correspondence among dimensional solutions, Pearson correlations were calculated between latent dimensions derived from 5-factor solutions derived from the autoencoder, EFA, PCA, and kernel PCA models. To account for multiple testing across correlation analyses, p-values were adjusted using the Benjamini–Hochberg false discovery rate procedure, with statistical significance defined as FDR-adjusted q < 0.05.

## Results

Parallel analyses tested factor solutions with dimensions up to 9 dimensions (supplementary Figure 1). Based on subsequent inspection of factor interpretability and pattern stability, the 3, 4, 5 solutions were retained for further comparison (supplementary Figure 2, 3, 4).

Across encoding dimensions 3 to 5, the autoencoder yielded the largest values of explained variance relative to all baseline methods, with its advantage remaining consistent at each evaluated dimensionality. Explained variance increased monotonically with higher encoding dimension for every method, but the autoencoder showed a fast rise, reaching approximately 0.42 at dimension 5. Among all selected kernels used for kernel PCA, the radial basis function kernel (RBF) kernel PCA provided the best performance, so only RBF kernel PCA is reported here. PCA and kernel PCA showed an intermediate range and across dimensions, with kernel PCA marginally higher than PCA at each setting. EFA produced the lowest explained variance throughout. Overall, the primary result is that the autoencoder achieves the highest total explained variance consistently across the examined encoding dimensions.

The alignment between the autoencoder derived latent space and established psychopathology dimensions was assessed by projecting each latent factor approximately onto the HiTOP spectra. Descriptively, largely distinct correspondence patterns were observed across the five latent factors, indicating that the learned representations captured differentiated configurations of psychopathological features rather than a single undifferentiated dimension (Figure 3). However, we also observed some overlap across factors, indicating that they do not reflect entirely distinct dimensions. Factor 1 was primarily characterized by antagonism and disinhibition, forming a latent factor aligned with externalizing behavioural disorders and showing weaker representation of internalizing and somatoform dimensions. In contrast, factor 2 reflected a mixed antagonism and internalizing dimension, capturing the existence of oppositional tendencies and internal distress, while showing comparatively limited contributions from thought disorder and disinhibition, and thereby differentiates antagonism accompanied by negative emotions from disinhibitory patterns. Factor 3 showed a dominant association with the somatoform dimension, with minimal contributions from other spectra, indicating a latent factor based on bodily and somatic symptom expression. Factors 4 and 5 captured mixed latent features. Factor 4 was also characterized by combined antagonism and internalizing dimensions, but represented a broader configuration of internalizing and antagonistic content than factor 2, reflecting a combination of oppositional conflict and negative emotion.

**Figure 1.**
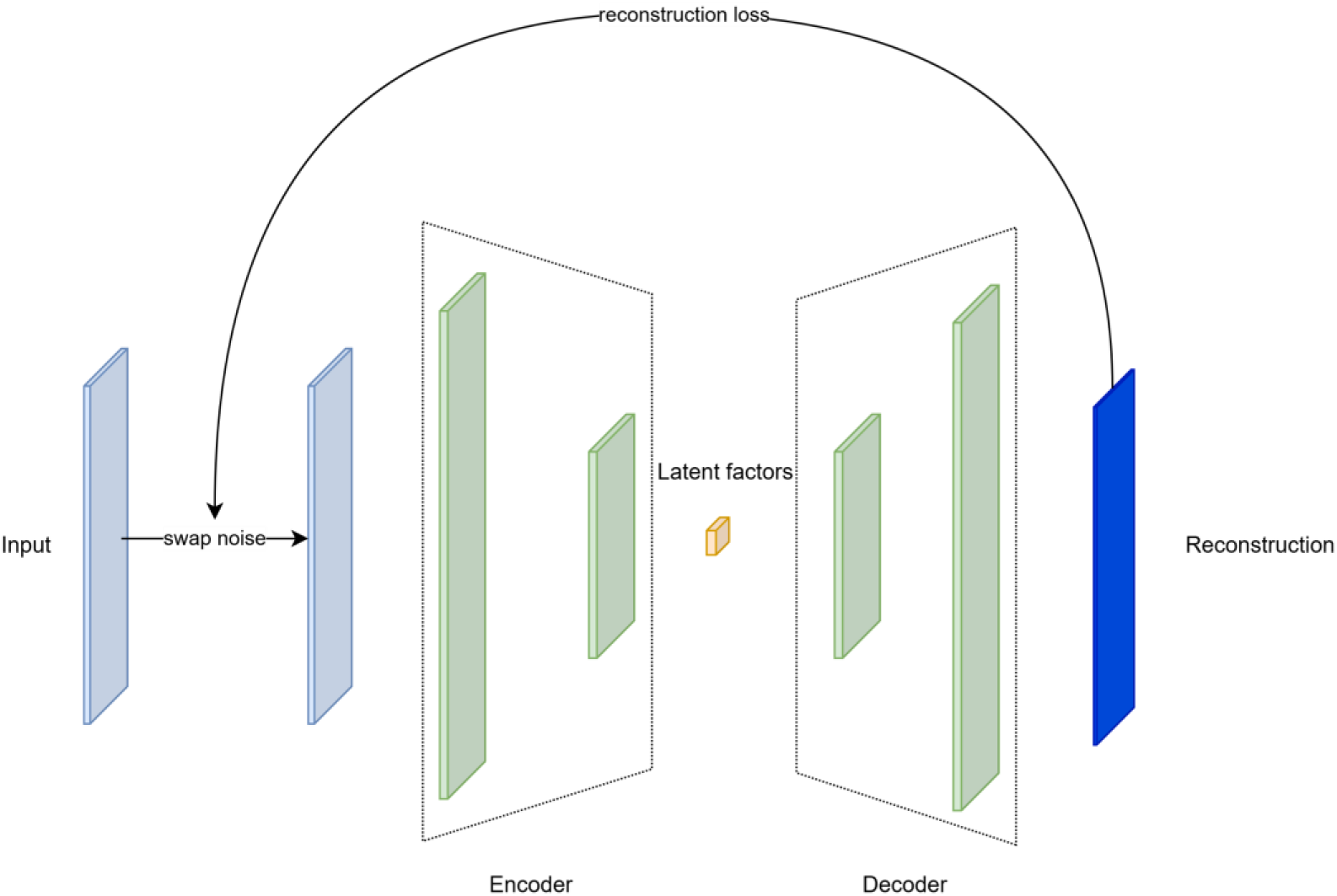
Architecture of the denoising autoencoder. The original input was corrupted using swap noise and then passed through the encoder to obtain a low-dimensional latent representation. The decoder reconstructed the original input from this latent representation, and the model was optimized by minimizing the reconstruction loss between the reconstructed output and the uncorrupted input.

**Figure 2.**
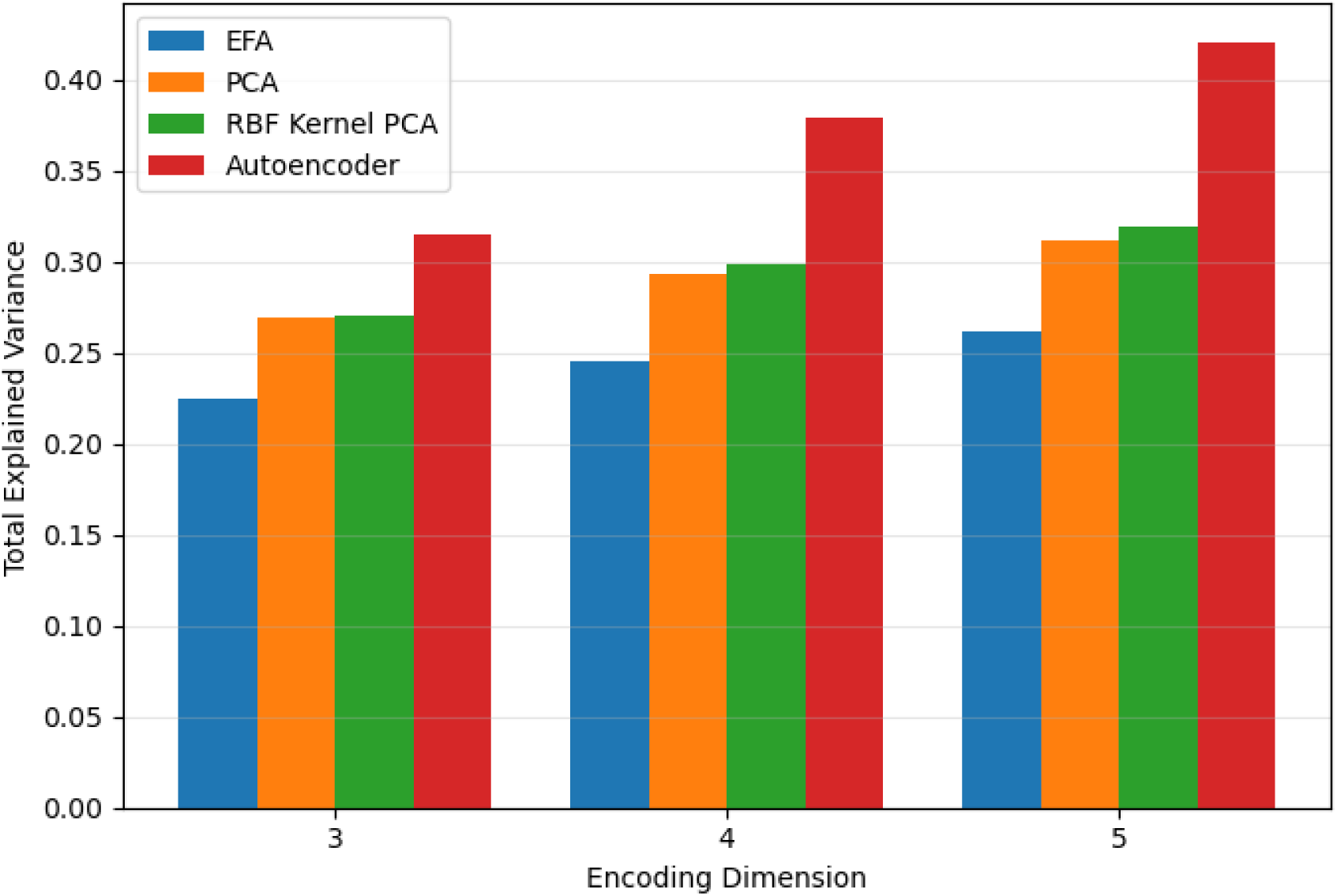
Total explained variance across encoding dimensions for different dimensionality reduction models. The bar chart compares the total explained variance obtained by EFA, PCA, RBF Kernel PCA, and the autoencoder when the encoding dimension was set to 3, 4, and 5. Across all encoding dimensions, the autoencoder achieved the highest total explained variance, indicating that the nonlinear autoencoder representation captured more variance in the high-dimensional symptom data than the linear and kernel-based alternatives.

**Figure 3.**
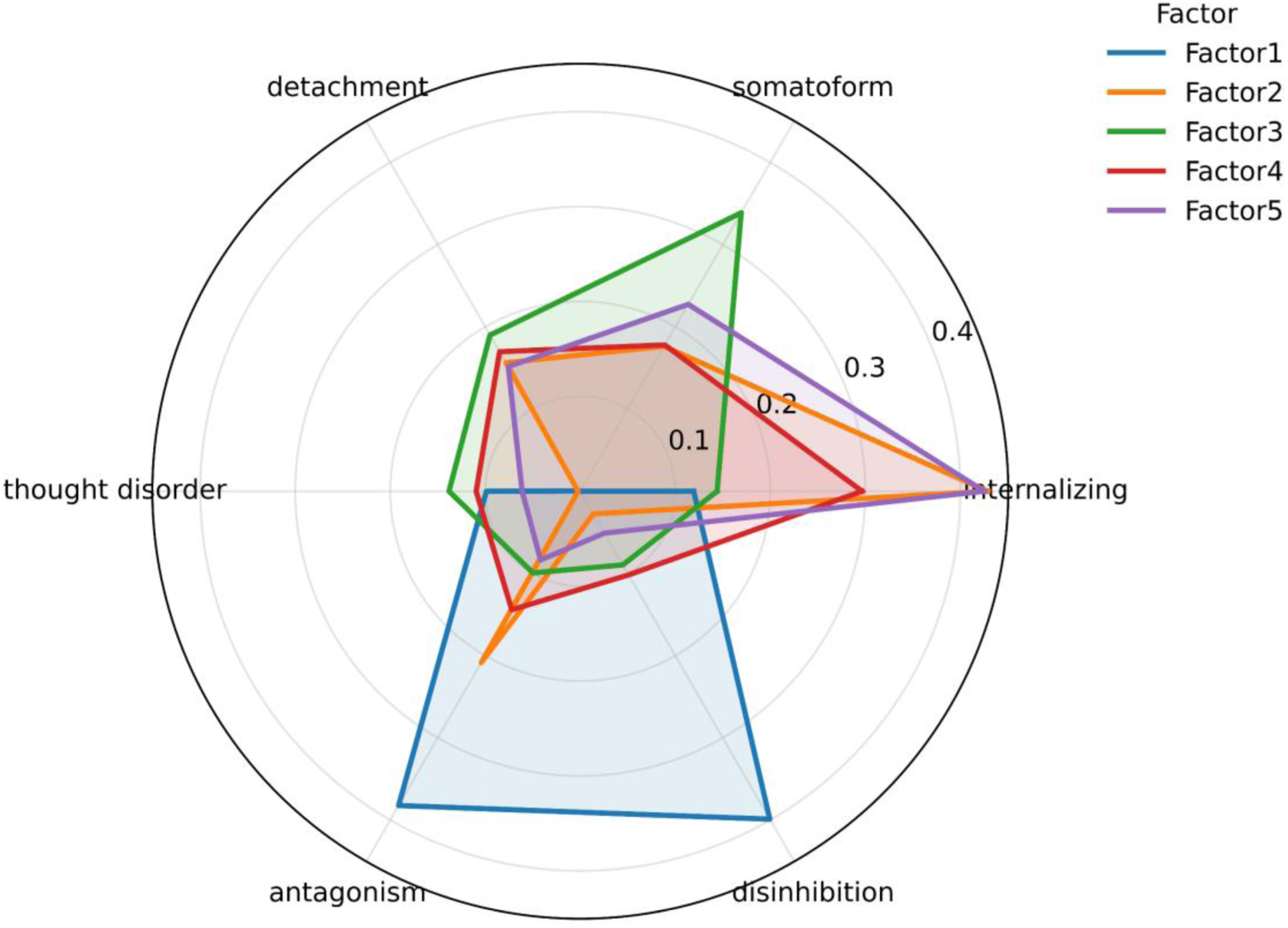
Radar chart for autoencoder derived 5 latent factors aligning with HiTOP spectra. The radar plot shows the relative alignment of the five autoencoder-derived latent factors with six HiTOP spectra: internalizing, somatoform, detachment, thought disorder, antagonism, and disinhibition. Each coloured shape represents one latent factor, and higher values indicate stronger correspondence with a given HiTOP spectrum.

Factor 5 was characterized by the somatoform and internalizing spectra, integrating bodily symptom expression with internal emotional distress. Together, these patterns demonstrate that the autoencoder latent space differentiated distinct configurations of psychopathological dimensions aligned with the HiTOP framework. These dimensions should therefore be interpreted as partially overlapping, data-driven representations with post-hoc correspondence to established psychopathology domains, rather than as discrete or one-to-one HiTOP spectra.

The cross-method correlation analysis showed partial convergence between autoencoder-derived dimensions and dimensions obtained from EFA, PCA, and kernel PCA. Although several autoencoder-derived dimensions were moderately to strongly associated with dimensions from the comparison methods, the overall pattern did not suggest a simple one-to-one mapping. This indicates that the autoencoder solution captured related but non-identical representations of adolescent mental health symptoms. Full correlation heatmaps are reported in Supplementary Figure 6.

### Predictive validity across external validators

To evaluate the predictive validity of the learned latent representations, we examined prediction performance on a held-out test set of external validators spanning mental health service use, medical history, medication use, family conflict, school connectedness, cognitive ability, academic performance, developmental delays, and peer relationships. Across all domains, predictive performance increased monotonically as a function of the number of latent factors, indicating that higher dimensional representations captured progressively more information relevant to downstream functional, clinical, and cognitive outcomes (Figure 4). This pattern was consistent with expectations from hierarchical models of childhood psychopathology (e.g., Michelini et al., 2019), in which broader dimensions account for substantial variance at low dimensionalities, with incremental gains achieved by higher dimensional factors.

**Figure 4.**
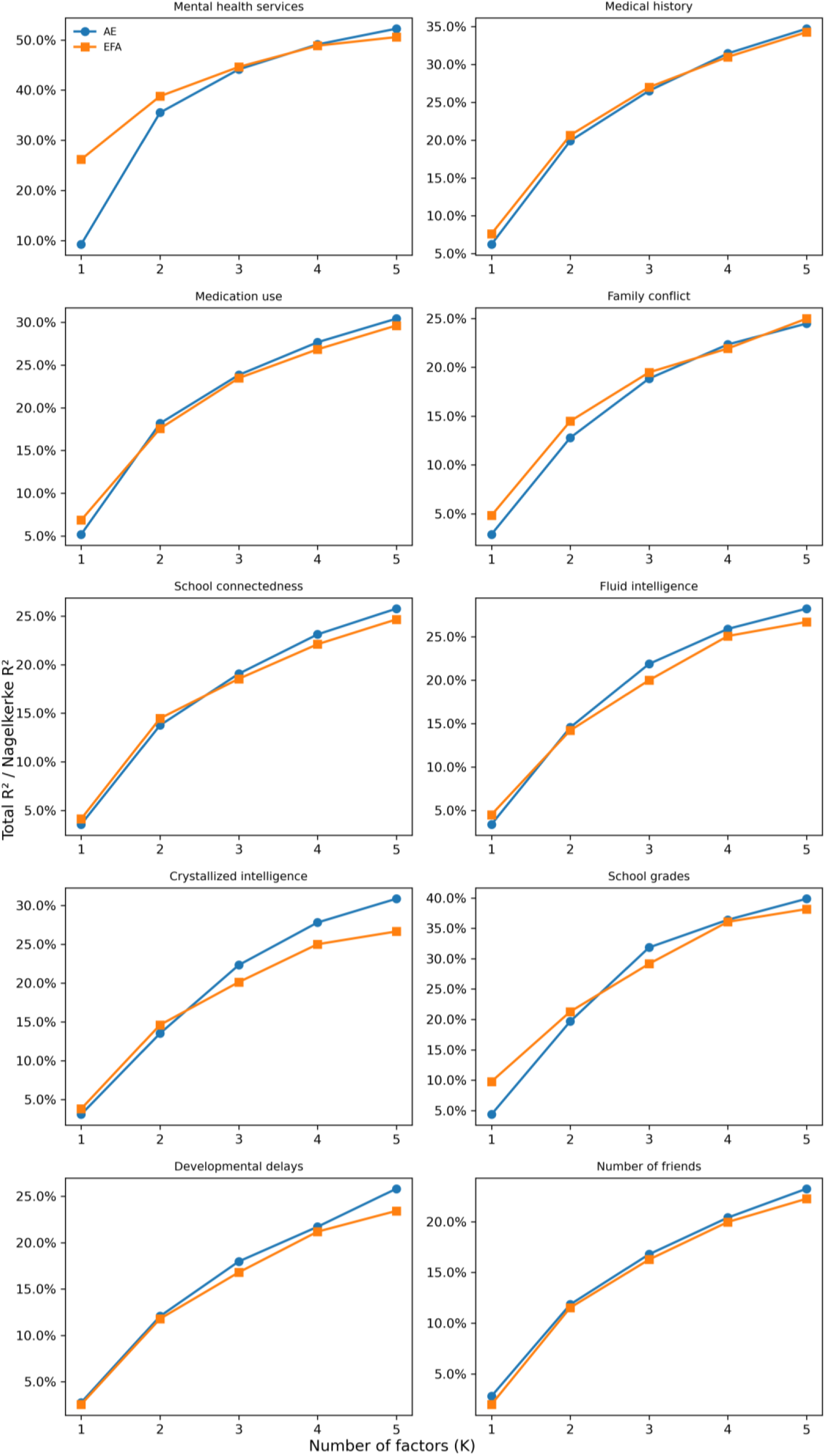
Cumulative explanatory power of autoencoder and EFA latent factors for validator outcomes. Cumulative explanatory power was evaluated across one to five latent factors using total R^2^ for continuous validators and Nagelkerke R^2^ for categorical validators. Both autoencoder- and EFA-derived factors showed increasing explanatory power as the number of factors increased, and no validator domain demonstrated consistent superiority of EFA over autoencoder across the range of dimensionalities examined.

Comparing representation learning approaches, autoencoder derived factors demonstrated equal or superior predictive validity relative to EFA across most validators, particularly at moderate to higher dimensionalities. The largest advantages of autoencoder were observed for cognitive and academic outcomes, including fluid intelligence, crystallized intelligence, and school grades, where autoencoder representations yielded consistently higher explained variance than EFA. For health service related and clinical history measures (e.g., mental health services use, medical history, medication use), both approaches showed comparable performance, with small but consistent gains for autoencoder at higher dimensions.

Importantly, no domain showed systematic superiority of EFA over autoencoder across dimensionalities, suggesting that the nonlinear representation learned by the autoencoder captured clinically and functionally meaningful structure beyond that obtained using linear factor models. Together, these results support the external validity of the learned latent dimensions and indicate that representation learning approaches provide a flexible and predictive framework for modelling hierarchical structure in childhood psychopathology.

## Discussion

This study evaluated whether denoising autoencoders could derive low-dimensional latent representations of youth psychopathology that are generalisable, interpretable, and externally valid. In this study, we addressed the need for more robust empirical evaluation of representation-learning approaches for transdiagnostic phenotyping via a denoising autoencoder and explicitly benchmarked this approach against commonly used linear and nonlinear alternatives within a held-out reconstruction and external-validation framework. Three main findings emerged. First, across the examined latent dimensionalities, the autoencoder achieved higher held-out reconstruction explained variance than PCA, EFA, and kernel PCA. Second, the learned dimensions showed interpretable item-level patterns, although these should be understood as data-driven, post-hoc mappings rather than direct one-to-one representations of HiTOP spectra or the original CBCL syndrome scales. Third, the autoencoder-derived dimensions showed external predictive validity across clinical, cognitive, family, school, developmental, and peer-related validators, with performance comparable to or modestly stronger than EFA across most domains.

Our findings suggest that representation learning with an autoencoder can capture structured, generalisable variance in youth symptom data that is not fully recovered by conventional linear factor models. One plausible explanation for the superior reconstruction performance is that the nonlinear encoder–decoder architecture can accommodate interactions, thresholds, and curved relationships among symptoms that violate linear factor assumptions, especially after ordinal-aware preprocessing. This interpretation is supported by prior work showing that autoencoders can represent complex structure in high-dimensional behavioural data without imposing orthogonality or simple covariance constraints (Bengio et al., 2013; Hinton & Salakhutdinov, 2006; Milano et al., 2024). Nonetheless, this substantial increase in explained variance within the questionnaire measures relative to linear approaches did not translate into an equal increase in variance explained within external validators, and future research could investigate the source of this discrepancy. For example, more directly testing how these subtle patterns within questionnaire responding may be used to predict external variances may be fruitful.

By training the autoencoder to reconstruct clean inputs from corrupted versions and by aggregating highly redundant items before model fitting, the learned latent space may emphasise stable, shared signal rather than item-specific noise. This could contribute to both improved generalisation and clearer alignment with broad psychopathology spectra, although the relative contribution of denoising versus architectural nonlinearity cannot be disentangled within the present design.

The differentiated HiTOP alignment across factors suggests that the learned representations capture more than a single broad dimension of psychopathology, but instead reflect configurations of higher-order dimensions, including mixed antagonism–internalizing and somatoform–internalizing patterns. This is consistent with hierarchical models proposing both broad general liability and partially independent spectra, rather than a single dominant dimension (Caspi et al., 2014; Kotov et al., 2017; Lahey et al., 2021).

Importantly, while these alignments support construct validity, they do not imply that the autoencoder directly estimates HiTOP dimensions; rather, the correspondence suggests convergent structure. The stronger predictive performance of autoencoder representations for cognitive and academic validators may indicate that nonlinear symptom embeddings can better capture behaviourally relevant variance that generalises beyond clinical service use or diagnostic proxies. This interpretation remains cautious, as prediction performance differences were modest and domain-specific, and causal mechanisms cannot be inferred from the present analyses.

The present findings are broadly consistent with the literature (Michelini et al., 2019) in showing monotonic gains in external validity with increasing dimensionality and in recovering dimensions aligned with antagonism, internalizing, and somatic symptom domains. A key contribution of this study is the explicit benchmarking of representation learning against conventional dimensionality reduction approaches under a held-out reconstruction framework, while using nested cross-validation and seed control to reduce optimism and underspecification. Prior applications of autoencoders in mental-health and health-record data have demonstrated their potential for phenotyping and prediction (Miotto et al., 2016), but often without direct comparison to factor models under equivalent evaluation conditions. The present results suggest that, when evaluated on generalisation rather than in-sample fit, autoencoders can recover representations that are at least as clinically meaningful as EFA while offering greater flexibility. The results do not imply that autoencoders replace factor analysis; rather, they highlight complementary inductive biases that may be advantageous for different inferential goals.

The findings also support the use of representation learning as a viable tool for constructing low-dimensional, generalisable symptom phenotypes in developmental mental health research, particularly when downstream validation and reproducibility are explicit goals. Autoencoder-derived latent factors may offer advantages for studies aiming to relate symptom structure to cognitive, educational, or biological measures, where nonlinear and interactive effects are plausible. However, these implications are supported only for the specific outcomes and dimensionalities examined here; broader clinical translation would require additional validation, including longitudinal prediction and replication in independent cohorts.

Our study has some limitations. All analyses relied on parent-reported CBCL data at a single baseline time point, limiting conclusions about developmental change or informant generalisability. This affects the temporal interpretation of latent structure and could be addressed with longitudinal modelling in future work. Moreover, the study focused on reconstruction EVR and external prediction as evaluation criteria; other notions of validity, such as sensitivity to rare symptoms or clinical decision thresholds, were not examined.

Future work could extend these findings in a number of ways. Replication using independent cohorts would test the stability of the learned representations over development. Including multi-informant data could test whether representation learning helps reconcile informant discrepancies. Linking latent factors to neuroimaging measures within the same framework would address mechanistic interpretability. These findings also have practical implications for researchers using the ABCD Study and similar large-scale developmental cohorts. The autoencoder-derived dimensions should not be viewed as replacements for the original CBCL syndrome scales or for EFA-derived factors. The original CBCL scales remain valuable when the goal is clinical interpretability, comparability with established norms, or alignment with prior CBCL-based research. EFA-derived factors remain useful when the aim is to estimate transparent, theory-aligned dimensions of psychopathology. In contrast, autoencoder-derived dimensions may be most useful when researchers aim to reduce high-dimensional item-level symptom data into compact representations that preserve out-of-sample reconstructive information and may capture non-linear patterns not fully represented by linear methods. Thus, the main implication is not that ABCD researchers should universally prefer autoencoder scores, but that these scores may provide a complementary representation for sensitivity analyses, predictive modelling, and studies focused on transdiagnostic symptom structure.

In summary, this study addresses the need for dimensionality reduction approaches in developmental psychopathology that consider the properties of questionnaire data while yielding generalisable and interpretable latent phenotypes. A further strength of this study is the use of a nested cross-validation framework with controlled random seeds, which provides a more rigorous assessment of model generalisability and reduces the risk that the reported findings are driven by a favourable data split or hyperparameter selection. By accounting for the ordinal nature of questionnaire data, redundancy reduction, and representation learning, and by benchmarking against established methods under rigorous evaluation, we show that autoencoder-derived representations can capture meaningful hierarchical structure and support external validity. These findings highlight both the promise and the boundaries of representation learning for mental health phenotypes, underscoring the importance of evaluation design and cautious interpretation. Continued work is needed to test robustness across time, informants, and biological domains.

## Data Availability

This study used data from the Adolescent Brain Cognitive Development (ABCD) Study, Curated Annual Release 5.1 (DOI: 10.15154/z563-zd24), held in the NIMH Data Archive (NDA). ABCD data are available to qualified researchers through application to the NDA (https://nda.nih.gov/abcd); the authors are not permitted to share individual-level data.

## Acknowledgements

Data used in the preparation of this article were obtained from the Adolescent Brain Cognitive Development (ABCD) Study (https://abcdstudy.org), held in the NIMH Data Archive (NDA). This is a multisite, longitudinal study designed to recruit more than 10,000 children age 9-10 and follow them over 10 years into early adulthood. A full list of supporters is available at https://abcdstudy.org/federal-partners.html. A listing of participating sites and a complete listing of the study investigators can be found at https://abcdstudy.org/consortium_members/. ABCD consortium investigators designed and implemented the study and/or provided data but did not necessarily participate in the analysis or writing of this report. This manuscript reflects the views of the authors and may not reflect the opinions or views of the NIH or ABCD consortium investigators. The ABCD data repository grows and changes over time. The ABCD data used in this report came from DOI: 10.15154/z563-zd24. DOIs can be found at https://nda.nih.gov/study.html?id=2313.

## Author contributions

LL was responsible for the main execution of the study, including data preprocessing, model development, statistical analysis, visualisation, interpretation of findings, and drafting of the manuscript. NC and GM contributed to the development of the research idea, provided methodological and conceptual guidance, and reviewed the manuscript critically. RI and TW supervised the project, advised on study design, methodology, and interpretation, and provided critical revisions. All authors read and approved the final manuscript.

## Code availability

Code to reproduce the analyses described here is available at https://github.com/ReveRoyl/ABCD_tabular_data_analysis.

## Supplementary Material

### Overview

This supplementary material provides additional information supporting the model selection, interpretability analyses, and item-level characterisation of the autoencoder-derived latent representations. Supplementary Figure 1 presents the reconstruction explained variance ratio across candidate latent dimensions and was used to inform the range of factor solutions examined in the main text. Supplementary Figures 2–4 show contour terrain maps of item contribution frequencies across canonical components for the three-, four-, and five-factor autoencoder solutions, respectively. Supplementary Table 1 provides the corresponding item-level loadings.

### Selection of latent dimensionality

Candidate autoencoder models were evaluated across latent dimensions up to 9 dimensions using reconstruction explained variance ratio. Latent dimensionality was not selected only based on reconstruction performance, because higher-dimensional solutions are expected to retain more information but may be less parsimonious and less interpretable. Therefore, the final dimensionality was considered using a combination of reconstruction performance, parsimony, and interpretability of factor profiles.

**Supplementary Figure 1.**
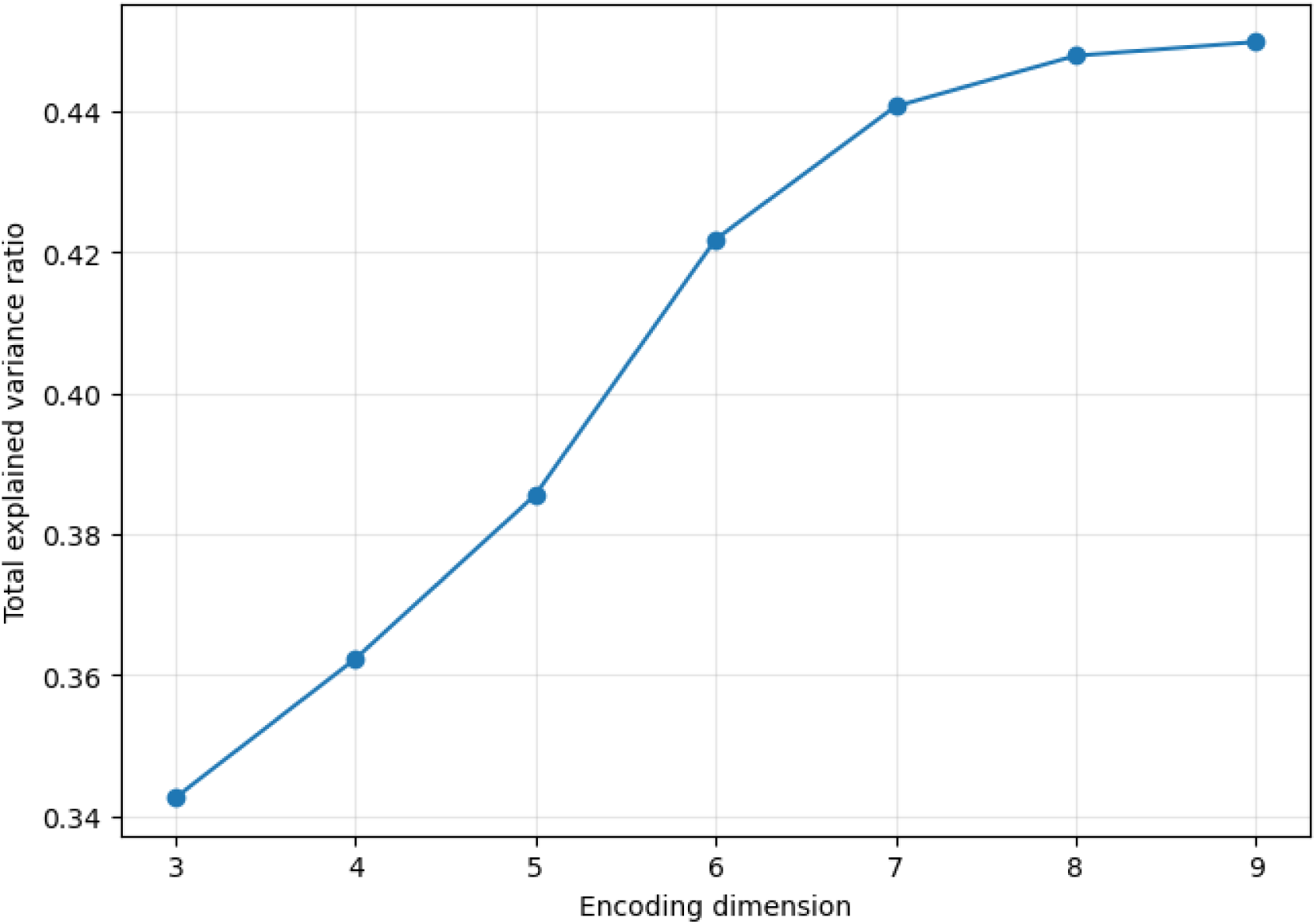
Autoencoder reconstruction explained variance ratio across latent dimensions.

This figure shows the reconstruction explained variance ratio of the autoencoder across candidate latent dimensions up to 9. Explained variance ratio increased as the latent dimensionality increased, indicating that higher-dimensional latent spaces retained more information from the original symptom features. The three-, four-, and five-factor solutions were selected for further inspection because they provided a balance between reconstruction performance, parsimony, and interpretability. The five-factor solution was retained as the primary solution in the main text because it showed strong reconstruction performance while allowing clinically interpretable differentiation across latent factors.

### Interpretability analysis

Interpretability was examined by relating autoencoder-derived latent factors to original symptom items using canonical correlation analysis. To assess the stability of item-level interpretability patterns, the autoencoder was trained 500 times using different random seeds, and item contribution patterns were summarised across these runs. Item-level contribution patterns were then visualised across canonical components using contour terrain maps. These maps were used to identify which CBCL items most consistently contributed to each canonical component and to support the post-hoc interpretation of the learned latent representations. The resulting factor labels should be interpreted as descriptive summaries of data-driven symptom configurations rather than as direct estimates of established diagnostic or HiTOP dimensions.

**Supplementary Figure 2.**
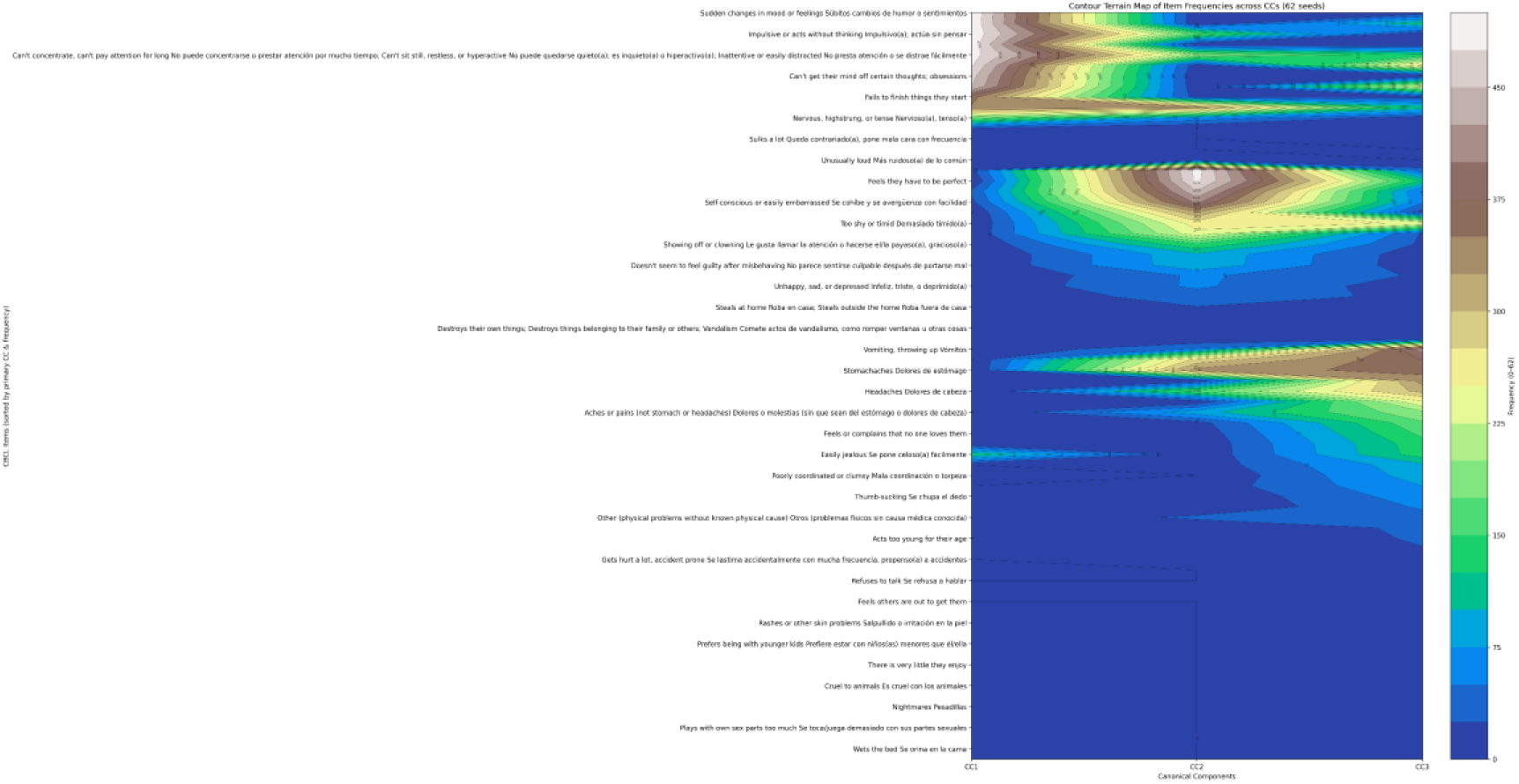
Contour terrain map of item frequencies across canonical components for the three-factor autoencoder solution.

**Supplementary Figure 3.**
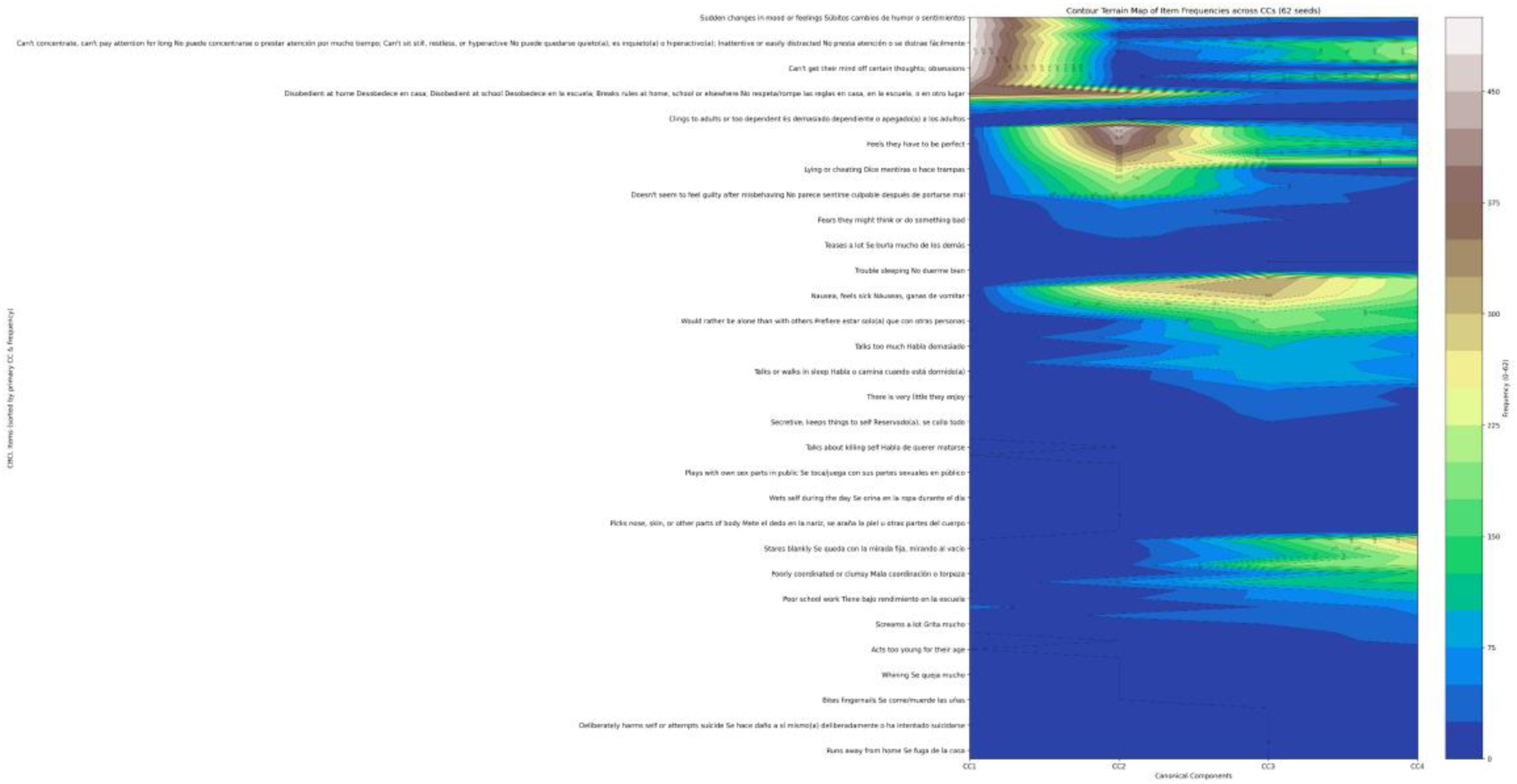
Contour terrain map of item frequencies across canonical components for the four-factor autoencoder solution.

**Supplementary Figure 4.**
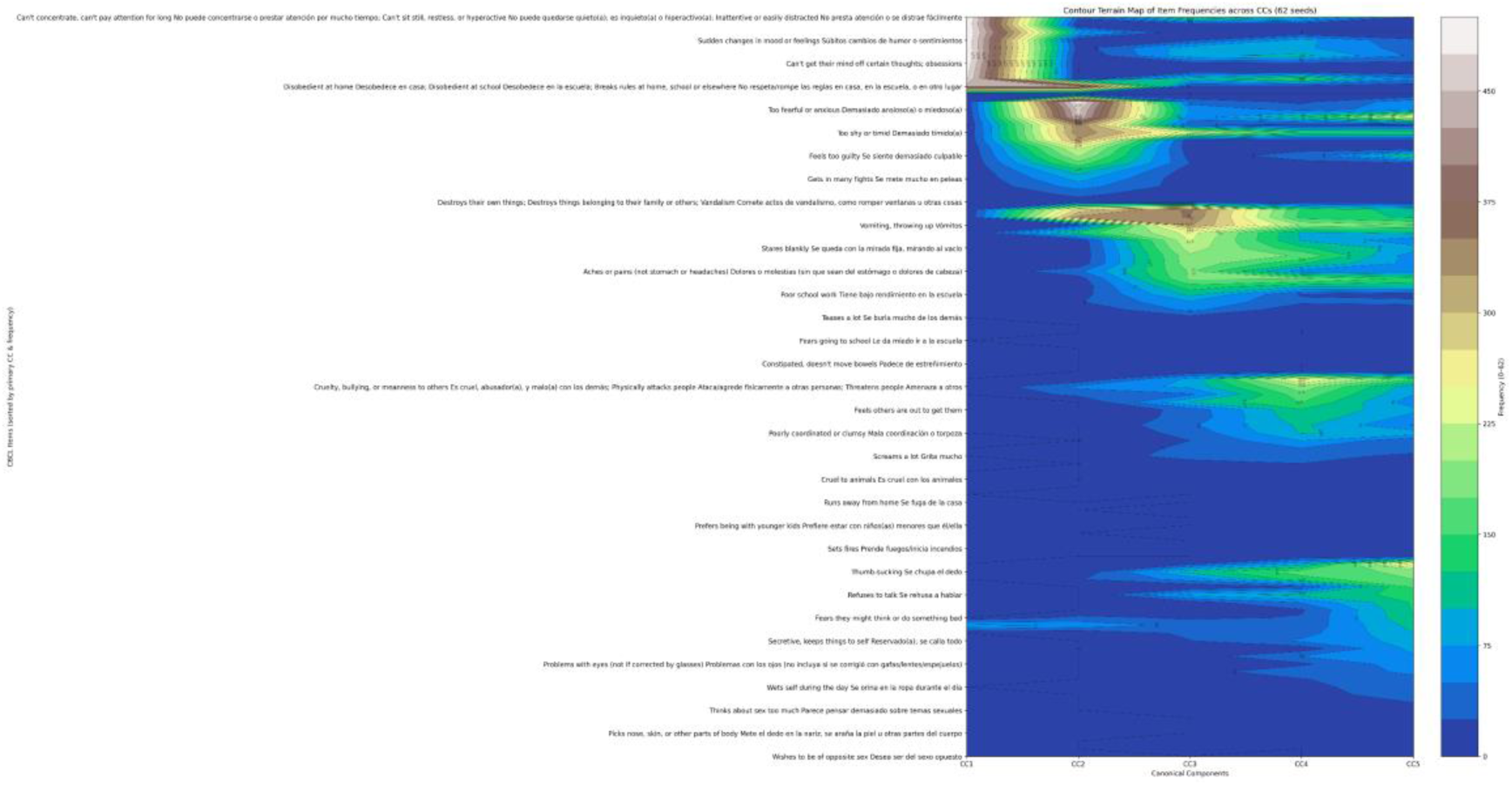
Contour terrain map of item frequencies across canonical components for the five-factor autoencoder solution.

**Supplementary Figure 5.**
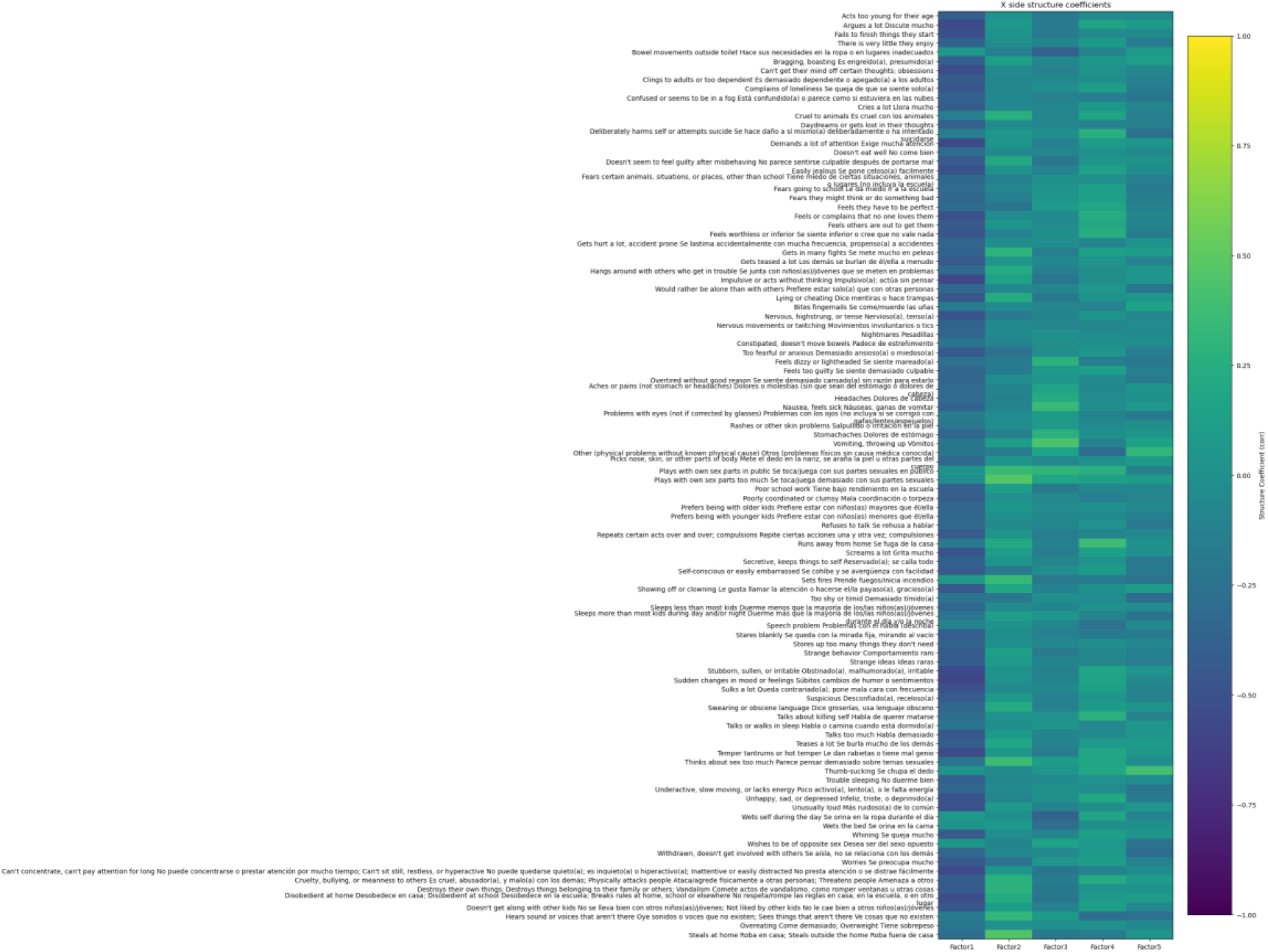
Item-level loading heatmap for the five-factor autoencoder solution.

This heatmap presents the item-level loading profiles of the original symptom features across the five autoencoder-derived latent factors. Loadings were obtained from the canonical correlation analysis between the latent factors and the original item-level features. Warmer and cooler values indicate stronger positive or negative associations, respectively. The heatmap was used to examine which symptom items contributed most strongly to each latent factor and to support the post-hoc interpretation of the learned representations.

Supplementary Table 1. Item-level loadings for the five-factor autoencoder solution.

Supplementary Table 1 is presented at the end of this supplementary material. This table reports the item-level loadings of the original symptom features on each of the five autoencoder-derived latent factors. Loadings were calculated as correlations between standardized observed features and canonical variates obtained from the canonical correlation analysis. Larger absolute values indicate stronger item-level contributions to the corresponding latent factor. The table is provided to support the interpretability of the five-factor solution and to allow readers to inspect the symptom content underlying each latent representation.

For the correspondence analysis, Pearson correlations were computed across participants between all latent dimensions derived from the autoencoder, EFA, PCA, and kernel PCA models. Statistical significance was assessed using two-sided tests, and p-values were adjusted for multiple comparisons using the Benjamini–Hochberg false discovery rate procedure.

**Supplementary Figure 6.**
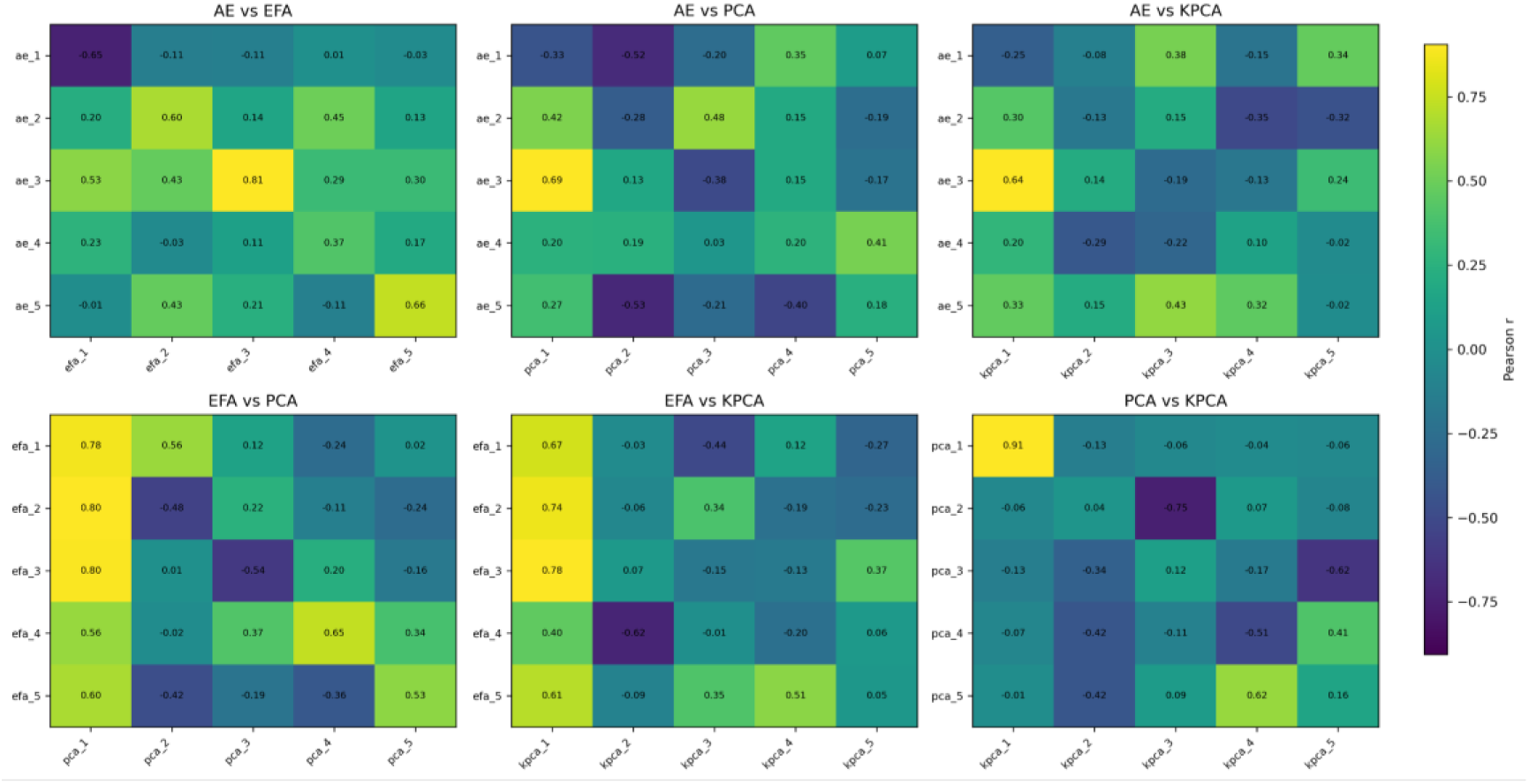
Pearson correlation heatmaps showing cross-method correspondence among five-dimensional representations derived from the autoencoder, EFA, PCA, and kernel PCA. Colours indicate the magnitude and direction of Pearson correlation coefficients. P-values were adjusted for multiple comparisons using the Benjamini–Hochberg false discovery rate procedure. All displayed correlations survived FDR correction at q < .05.

**Supplementary Table 1.**
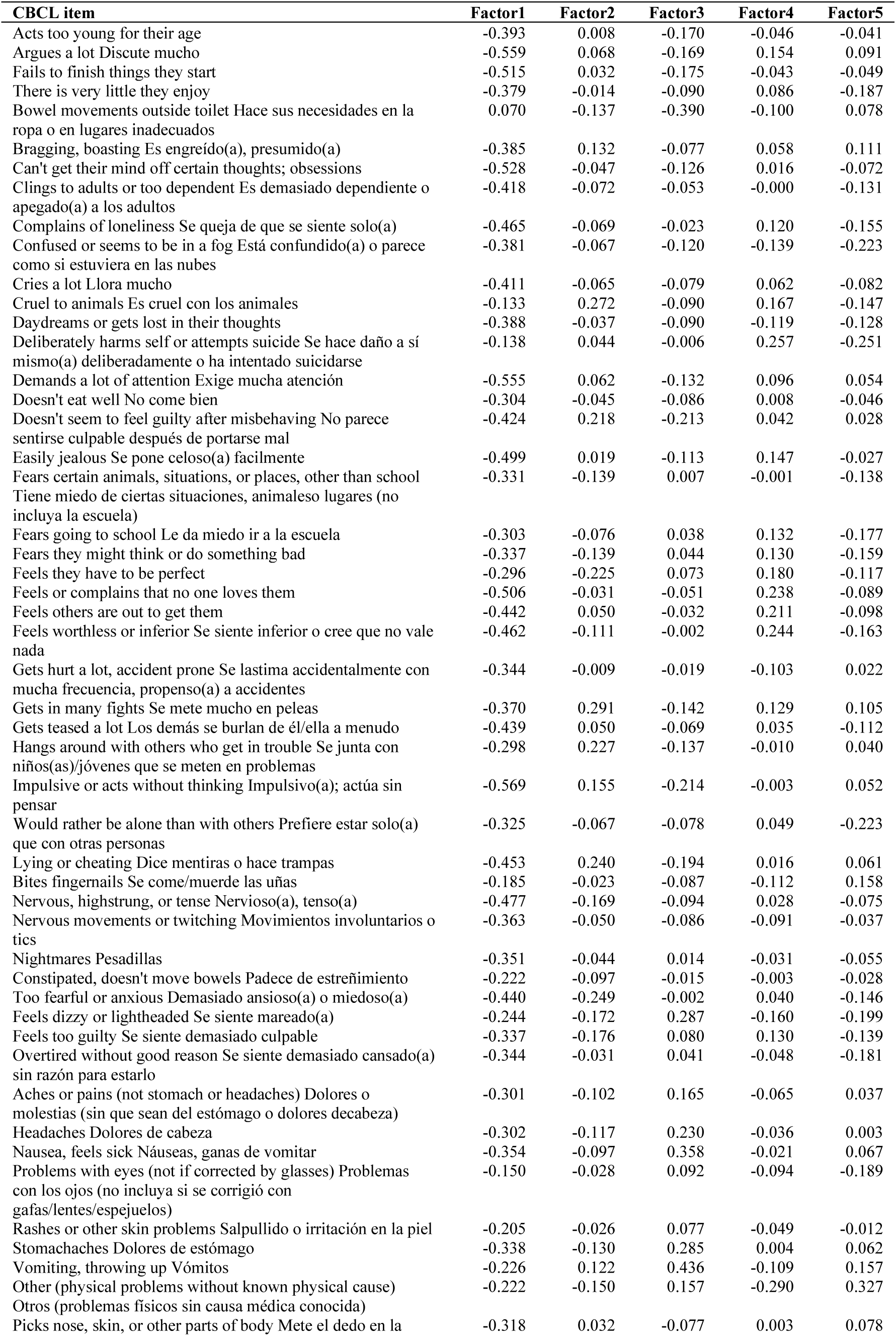
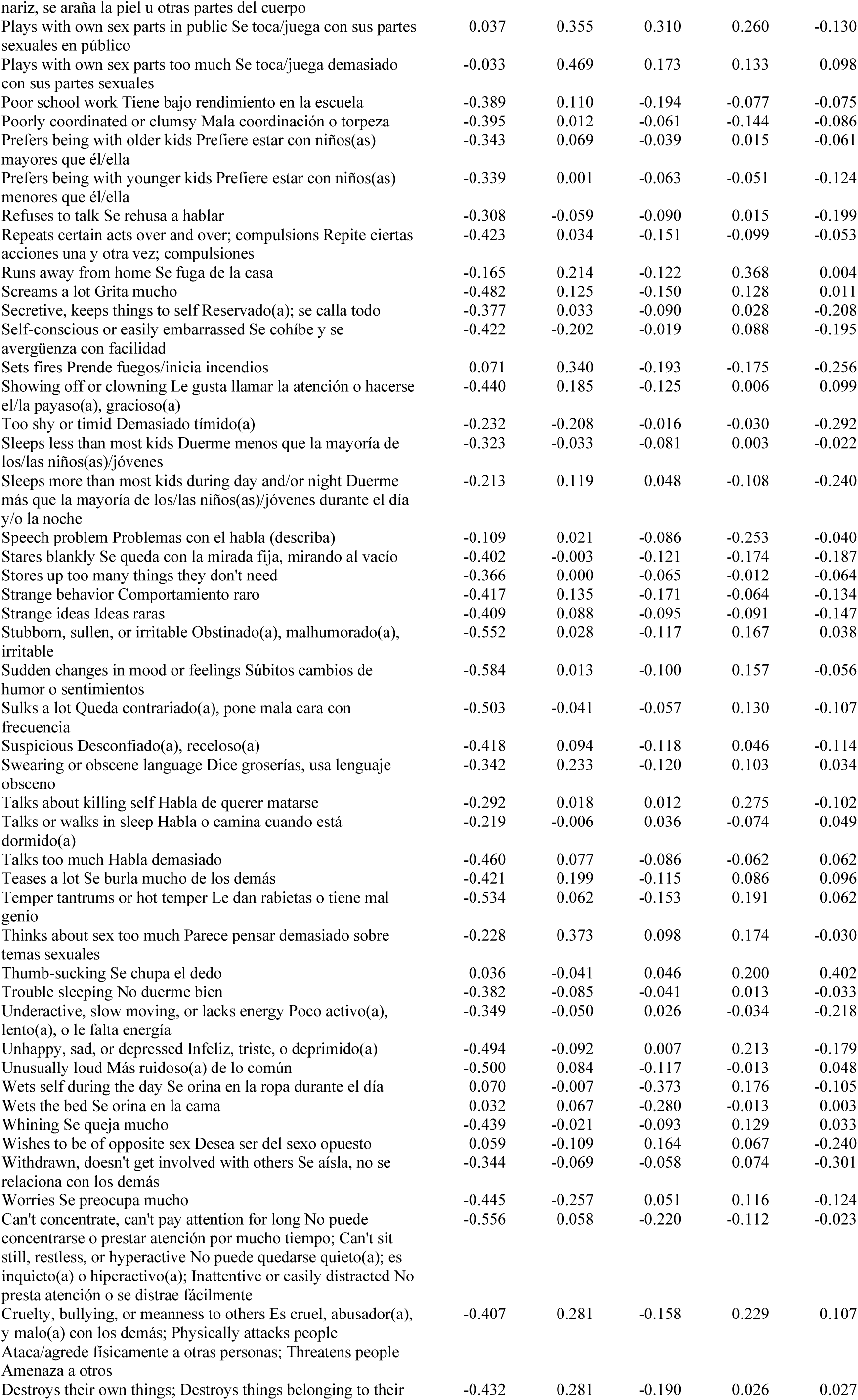
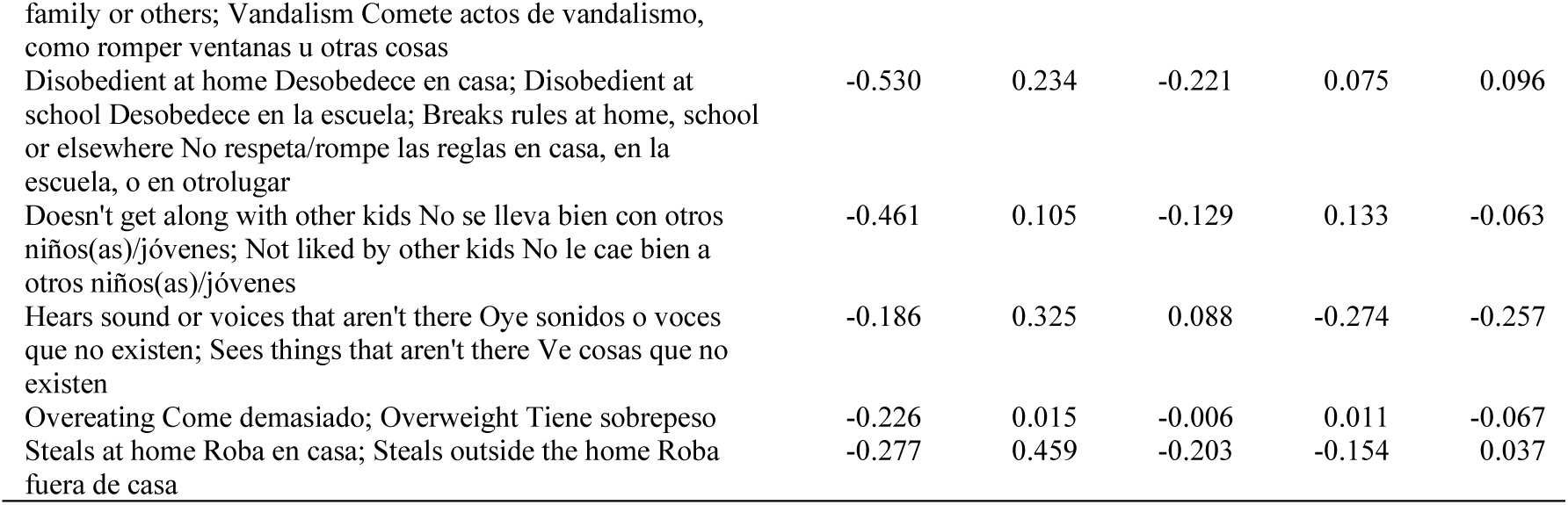
Item-level loadings for the five-factor autoencoder solution. Values are correlations between standardized CBCL item-level features and the canonical variates for each latent factor.

| **CBCL item** | **Factor1** | **Factor2** | **Factor3** | **Factor4** | **Factor5** |
|---|---|---|---|---|---|
| Acts too young for their age | -0.393 | 0.008 | -0.170 | -0.046 | -0.041 |
| Argues a lot Discute mucho | -0.559 | 0.068 | -0.169 | 0.154 | 0.091 |
| Fails to finish things they start | -0.515 | 0.032 | -0.175 | -0.043 | -0.049 |
| There is very little they enjoy | -0.379 | -0.014 | -0.090 | 0.086 | -0.187 |
| Bowel movements outside toilet Hace sus necesidades en la ropa o en lugares inadecuados | 0.070 | -0.137 | -0.390 | -0.100 | 0.078 |
| Bragging, boasting Es engreído(a), presumido(a) | -0.385 | 0.132 | -0.077 | 0.058 | 0.111 |
| Can't get their mind off certain thoughts; obsessions | -0.528 | -0.047 | -0.126 | 0.016 | -0.072 |
| Clings to adults or too dependent Es demasiado dependiente o apegado(a) a los adultos | -0.418 | -0.072 | -0.053 | -0.000 | -0.131 |
| Complains of loneliness Se queja de que se siente solo(a) | -0.465 | -0.069 | -0.023 | 0.120 | -0.155 |
| Confused or seems to be in a fog Está confundido(a) o parece como si estuviera en las nubes | -0.381 | -0.067 | -0.120 | -0.139 | -0.223 |
| Cries a lot Llora mucho | -0.411 | -0.065 | -0.079 | 0.062 | -0.082 |
| Cruel to animals Es cruel con los animales | -0.133 | 0.272 | -0.090 | 0.167 | -0.147 |
| Daydreams or gets lost in their thoughts | -0.388 | -0.037 | -0.090 | -0.119 | -0.128 |
| Deliberately harms self or attempts suicide Se hace daño a sí mismo(a) deliberadamente o ha intentado suicidarse | -0.138 | 0.044 | -0.006 | 0.257 | -0.251 |
| Demands a lot of attention Exige mucha atención | -0.555 | 0.062 | -0.132 | 0.096 | 0.054 |
| Doesn't eat well No come bien | -0.304 | -0.045 | -0.086 | 0.008 | -0.046 |
| Doesn't seem to feel guilty after misbehaving No parece sentirse culpable después de portarse mal | -0.424 | 0.218 | -0.213 | 0.042 | 0.028 |
| Easily jealous Se pone celoso(a) facilmente | -0.499 | 0.019 | -0.113 | 0.147 | -0.027 |
| Fears certain animals, situations, or places, other than school Tiene miedo de ciertas situaciones, animaleso lugares (no incluya la escuela) | -0.331 | -0.139 | 0.007 | -0.001 | -0.138 |
| Fears going to school Le da miedo ir a la escuela | -0.303 | -0.076 | 0.038 | 0.132 | -0.177 |
| Fears they might think or do something bad | -0.337 | -0.139 | 0.044 | 0.130 | -0.159 |
| Feels they have to be perfect | -0.296 | -0.225 | 0.073 | 0.180 | -0.117 |
| Feels or complains that no one loves them | -0.506 | -0.031 | -0.051 | 0.238 | -0.089 |
| Feels others are out to get them | -0.442 | 0.050 | -0.032 | 0.211 | -0.098 |
| Feels worthless or inferior Se siente inferior o cree que no vale nada | -0.462 | -0.111 | -0.002 | 0.244 | -0.163 |
| Gets hurt a lot, accident prone Se lastima accidentalmente con mucha frecuencia, propenso(a) a accidentes | -0.344 | -0.009 | -0.019 | -0.103 | 0.022 |
| Gets in many fights Se mete mucho en peleas | -0.370 | 0.291 | -0.142 | 0.129 | 0.105 |
| Gets teased a lot Los demás se burlan de él/ella a menudo | -0.439 | 0.050 | -0.069 | 0.035 | -0.112 |
| Hangs around with others who get in trouble Se junta con niños(as)/jóvenes que se meten en problemas | -0.298 | 0.227 | -0.137 | -0.010 | 0.040 |
| Impulsive or acts without thinking Impulsivo(a); actúa sin pensar | -0.569 | 0.155 | -0.214 | -0.003 | 0.052 |
| Would rather be alone than with others Prefiere estar solo(a) que con otras personas | -0.325 | -0.067 | -0.078 | 0.049 | -0.223 |
| Lying or cheating Dice mentiras o hace trampas | -0.453 | 0.240 | -0.194 | 0.016 | 0.061 |
| Bites fingernails Se come/muerde las uñas | -0.185 | -0.023 | -0.087 | -0.112 | 0.158 |
| Nervous, highstrung, or tense Nervioso(a), tenso(a) | -0.477 | -0.169 | -0.094 | 0.028 | -0.075 |
| Nervous movements or twitching Movimientos involuntarios o tics | -0.363 | -0.050 | -0.086 | -0.091 | -0.037 |
| Nightmares Pesadillas | -0.351 | -0.044 | 0.014 | -0.031 | -0.055 |
| Constipated, doesn't move bowels Padece de estreñimiento | -0.222 | -0.097 | -0.015 | -0.003 | -0.028 |
| Too fearful or anxious Demasiado ansioso(a) o miedoso(a) | -0.440 | -0.249 | -0.002 | 0.040 | -0.146 |
| Feels dizzy or lightheaded Se siente mareado(a) | -0.244 | -0.172 | 0.287 | -0.160 | -0.199 |
| Feels too guilty Se siente demasiado culpable | -0.337 | -0.176 | 0.080 | 0.130 | -0.139 |
| Overtired without good reason Se siente demasiado cansado(a) sin razón para estarlo | -0.344 | -0.031 | 0.041 | -0.048 | -0.181 |
| Aches or pains (not stomach or headaches) Dolores o molestias (sin que sean del estómago o dolores decabeza) | -0.301 | -0.102 | 0.165 | -0.065 | 0.037 |
| Headaches Dolores de cabeza | -0.302 | -0.117 | 0.230 | -0.036 | 0.003 |
| Nausea, feels sick Náuseas, ganas de vomitar | -0.354 | -0.097 | 0.358 | -0.021 | 0.067 |
| Problems with eyes (not if corrected by glasses) Problemas con los ojos (no incluya si se corrigió con gafas/lentes/espejuelos) | -0.150 | -0.028 | 0.092 | -0.094 | -0.189 |
| Rashes or other skin problems Salpullido o irritación en la piel | -0.205 | -0.026 | 0.077 | -0.049 | -0.012 |
| Stomachaches Dolores de estómago | -0.338 | -0.130 | 0.285 | 0.004 | 0.062 |
| Vomiting, throwing up Vómitos | -0.226 | 0.122 | 0.436 | -0.109 | 0.157 |
| Other (physical problems without known physical cause) Otros (problemas físicos sin causa médica conocida) | -0.222 | -0.150 | 0.157 | -0.290 | 0.327 |
| Picks nose, skin, or other parts of body Mete el dedo en la nariz, se araña la piel u otras partes del cuerpo | -0.318 | 0.032 | -0.077 | 0.003 | 0.078 |
| Plays with own sex parts in public Se toca/juega con sus partes sexuales en público | 0.037 | 0.355 | 0.310 | 0.260 | -0.130 |
| Plays with own sex parts too much Se toca/juega demasiado con sus partes sexuales | -0.033 | 0.469 | 0.173 | 0.133 | 0.098 |
| Poor school work Tiene bajo rendimiento en la escuela | -0.389 | 0.110 | -0.194 | -0.077 | -0.075 |
| Poorly coordinated or clumsy Mala coordinación o torpeza | -0.395 | 0.012 | -0.061 | -0.144 | -0.086 |
| Prefers being with older kids Prefiere estar con niños(as) mayores que él/ella | -0.343 | 0.069 | -0.039 | 0.015 | -0.061 |
| Prefers being with younger kids Prefiere estar con niños(as) menores que él/ella | -0.339 | 0.001 | -0.063 | -0.051 | -0.124 |
| Refuses to talk Se rehusa a hablar | -0.308 | -0.059 | -0.090 | 0.015 | -0.199 |
| Repeats certain acts over and over; compulsions Repite ciertas acciones una y otra vez; compulsiones | -0.423 | 0.034 | -0.151 | -0.099 | -0.053 |
| Runs away from home Se fuga de la casa | -0.165 | 0.214 | -0.122 | 0.368 | 0.004 |
| Screams a lot Grita mucho | -0.482 | 0.125 | -0.150 | 0.128 | 0.011 |
| Secretive, keeps things to self Reservado(a); se calla todo | -0.377 | 0.033 | -0.090 | 0.028 | -0.208 |
| Self-conscious or easily embarrassed Se cohíbe y se avergüenza con facilidad | -0.422 | -0.202 | -0.019 | 0.088 | -0.195 |
| Sets fires Prende fuegos/inicia incendios | 0.071 | 0.340 | -0.193 | -0.175 | -0.256 |
| Showing off or clowning Le gusta llamar la atención o hacerse el/la payaso(a), gracioso(a) | -0.440 | 0.185 | -0.125 | 0.006 | 0.099 |
| Too shy or timid Demasiado tímido(a) | -0.232 | -0.208 | -0.016 | -0.030 | -0.292 |
| Sleeps less than most kids Duerme menos que la mayoría de los/las niños(as)/jóvenes | -0.323 | -0.033 | -0.081 | 0.003 | -0.022 |
| Sleeps more than most kids during day and/or night Duerme más que la mayoría de los/las niños(as)/jóvenes durante el día y/o la noche | -0.213 | 0.119 | 0.048 | -0.108 | -0.240 |
| Speech problem Problemas con el habla (describa) | -0.109 | 0.021 | -0.086 | -0.253 | -0.040 |
| Stares blankly Se queda con la mirada fija, mirando al vacío | -0.402 | -0.003 | -0.121 | -0.174 | -0.187 |
| Stores up too many things they don't need | -0.366 | 0.000 | -0.065 | -0.012 | -0.064 |
| Strange behavior Comportamiento raro | -0.417 | 0.135 | -0.171 | -0.064 | -0.134 |
| Strange ideas Ideas raras | -0.409 | 0.088 | -0.095 | -0.091 | -0.147 |
| Stubborn, sullen, or irritable Obstinado(a), malhumorado(a), irritable | -0.552 | 0.028 | -0.117 | 0.167 | 0.038 |
| Sudden changes in mood or feelings Súbitos cambios de humor o sentimientos | -0.584 | 0.013 | -0.100 | 0.157 | -0.056 |
| Sulks a lot Queda contrariado(a), pone mala cara con frecuencia | -0.503 | -0.041 | -0.057 | 0.130 | -0.107 |
| Suspicious Desconfiado(a), receloso(a) | -0.418 | 0.094 | -0.118 | 0.046 | -0.114 |
| Swearing or obscene language Dice groserías, usa lenguaje obsceno | -0.342 | 0.233 | -0.120 | 0.103 | 0.034 |
| Talks about killing self Habla de querer matarse | -0.292 | 0.018 | 0.012 | 0.275 | -0.102 |
| Talks or walks in sleep Habla o camina cuando está dormido(a) | -0.219 | -0.006 | 0.036 | -0.074 | 0.049 |
| Talks too much Habla demasiado | -0.460 | 0.077 | -0.086 | -0.062 | 0.062 |
| Teases a lot Se burla mucho de los demás | -0.421 | 0.199 | -0.115 | 0.086 | 0.096 |
| Temper tantrums or hot temper Le dan rabietas o tiene mal genio | -0.534 | 0.062 | -0.153 | 0.191 | 0.062 |
| Thinks about sex too much Parece pensar demasiado sobre temas sexuales | -0.228 | 0.373 | 0.098 | 0.174 | -0.030 |
| Thumb-sucking Se chupa el dedo | 0.036 | -0.041 | 0.046 | 0.200 | 0.402 |
| Trouble sleeping No duerme bien | -0.382 | -0.085 | -0.041 | 0.013 | -0.033 |
| Underactive, slow moving, or lacks energy Poco activo(a), lento(a), o le falta energía | -0.349 | -0.050 | 0.026 | -0.034 | -0.218 |
| Unhappy, sad, or depressed Infeliz, triste, o deprimido(a) | -0.494 | -0.092 | 0.007 | 0.213 | -0.179 |
| Unusually loud Más ruidoso(a) de lo común | -0.500 | 0.084 | -0.117 | -0.013 | 0.048 |
| Wets self during the day Se orina en la ropa durante el día | 0.070 | -0.007 | -0.373 | 0.176 | -0.105 |
| Wets the bed Se orina en la cama | 0.032 | 0.067 | -0.280 | -0.013 | 0.003 |
| Whining Se queja mucho | -0.439 | -0.021 | -0.093 | 0.129 | 0.033 |
| Wishes to be of opposite sex Desea ser del sexo opuesto | 0.059 | -0.109 | 0.164 | 0.067 | -0.240 |
| Withdrawn, doesn't get involved with others Se aísla, no se relaciona con los demás | -0.344 | -0.069 | -0.058 | 0.074 | -0.301 |
| Worries Se preocupa mucho | -0.445 | -0.257 | 0.051 | 0.116 | -0.124 |
| Can't concentrate, can't pay attention for long No puede concentrarse o prestar atención por mucho tiempo; Can't sit still, restless, or hyperactive No puede quedarse quieto(a); es inquieto(a) o hiperactivo(a); Inattentive or easily distracted No presta atención o se distrae fácilmente | -0.556 | 0.058 | -0.220 | -0.112 | -0.023 |
| Cruelty, bullying, or meanness to others Es cruel, abusador(a), y malo(a) con los demás; Physically attacks people Ataca/agrede físicamente a otras personas; Threatens people Amenaza a otros | -0.407 | 0.281 | -0.158 | 0.229 | 0.107 |
| Destroys their own things; Destroys things belonging to their family or others; Vandalism Comete actos de vandalismo, como romper ventanas u otras cosas | -0.432 | 0.281 | -0.190 | 0.026 | 0.027 |
| Disobedient at home Desobedece en casa; Disobedient at school Desobedece en la escuela; Breaks rules at home, school or elsewhere No respeta/rompe las reglas en casa, en la escuela, o en otrolugar | -0.530 | 0.234 | -0.221 | 0.075 | 0.096 |
| Doesn't get along with other kids No se lleva bien con otros niños(as)/jóvenes; Not liked by other kids No le cae bien a otros niños(as)/jóvenes | -0.461 | 0.105 | -0.129 | 0.133 | -0.063 |
| Hears sound or voices that aren't there Oye sonidos o voces que no existen; Sees things that aren't there Ve cosas que no existen | -0.186 | 0.325 | 0.088 | -0.274 | -0.257 |
| Overeating Come demasiado; Overweight Tiene sobrepeso | -0.226 | 0.015 | -0.006 | 0.011 | -0.067 |
| Steals at home Roba en casa; Steals outside the home Roba fuera de casa | -0.277 | 0.459 | -0.203 | -0.154 | 0.037 |

